# A CBC-derived score associated with dengue NS1 antigen positivity among suspected dengue patients in Dhaka: a retrospective model-development study

**DOI:** 10.64898/2026.11.05.26367350

**Authors:** Rasel Khan Nishan, Md. Shaha Alam, Abul Kalam Azad, Md. Murshed Hasan Sarkar, Tanjum Ahmed Nodee, Md. Zakaria Mia, Md. Ibrahim Miah, Md. Shariful Islam

## Abstract

**Background:** Early diagnostic triage for suspected dengue is important in high-volume settings, yet confirmatory testing may be delayed or intermittently available. This study aimed to develop and internally validate a simple complete blood count (CBC)-derived score to estimate the likelihood of same-day dengue non-structural protein 1 (NS1) antigen positivity among suspected dengue patients in Dhaka, Bangladesh.

**Methods:** We conducted a retrospective model-development study at Uttara Adhunik Medical College Hospital using data from patients with clinical suspicion of dengue between 19 August and 12 October 2023. The final analysis included 596 unique patients who underwent CBC and NS1 testing on the same day. Least absolute shrinkage and selection operator (LASSO) penalized logistic regression was used for variable selection among routinely available demographic and hematological predictors. Selected predictors were refit using unpenalized logistic regression to derive a CBC score. Internal validation and calibration were assessed using bootstrap optimism correction with 400 resamples.

**Results:** Among 596 patients, 306 (51.3%) were NS1 positive. The final parsimonious model retained platelet count rescaled per 10 x 10^9^/L, white blood cell count, and monocyte count. The optimism-corrected AUC was 0.9429. At the Youden-derived cutoff of 11.825, the final integer score had sensitivity 0.833 (95% CI, 0.787-0.873), specificity 0.966 (95% CI, 0.938-0.983), PPV 0.962 (95% CI, 0.932-0.982), NPV 0.846 (95% CI, 0.802-0.883), and accuracy 0.898 (95% CI, 0.870-0.921). A rounded cutoff of 12 produced similar performance. In benchmark analysis, the final model showed a modest improvement over platelet count alone (AUC 0.944 vs 0.931; Delta AUC 0.012; DeLong p=0.00364).

**Conclusion:** A three-parameter CBC score can estimate the likelihood of NS1 antigen positivity among suspected dengue patients in this single-center dataset. The score should be interpreted as a CBC-only triage adjunct, not as a replacement for confirmatory dengue testing. External validation in independent cohorts is needed before routine clinical use.

## Introduction

Dengue is a rapidly expanding global public-health problem. WHO reports that dengue incidence has increased markedly in recent decades, with 2024 recording historically high reported case numbers and more than 100 countries affected [1,2]. The burden is especially relevant in tropical urban settings where Aedes mosquito exposure, population density, and seasonal outbreaks can produce large surges of febrile illness [1-3]. Bangladesh has experienced major recent dengue outbreaks. DGHS reported 321,179 dengue cases and 1,705 deaths in 2023, followed by 101,214 cases and 575 deaths in 2024 [4,5]. DGHS dashboard reporting and WHO outbreak documentation further emphasize the operational pressure created by recurrent dengue admissions and deaths in Bangladesh [6,7]. During outbreak periods, clinicians in Dhaka often need to prioritize testing and follow-up while managing many patients with overlapping febrile presentations.

Dengue diagnosis depends on illness phase and test availability. Virologic approaches, including NS1 antigen testing and nucleic-acid amplification tests, are most useful during the acute phase, whereas serology becomes more informative later [8-10]. NS1 testing is widely used, but a negative NS1 result does not rule out dengue infection, and performance varies by timing of sampling, serotype, and immune status [9,10,30-32]. Therefore, routine laboratory patterns should not replace confirmatory testing, but they may help triage suspected cases when testing is delayed or constrained.

CBC parameters are inexpensive, widely available, and commonly ordered for suspected dengue. Prior studies have repeatedly identified leukopenia and thrombocytopenia as useful markers in dengue assessment [11]. Tuan and colleagues developed an early dengue classifier that incorporated age, white blood cell count, and platelet count [12]. Poottasane and colleagues also reported a CBC-based score including white blood cell count, platelet count, and monocyte distribution width, while emphasizing the need for local validation [13].

Despite this evidence, there remains a need for simple locally derived tools that use only routinely available parameters in Bangladeshi outbreak settings. We therefore aimed to develop and internally validate a parsimonious CBC-derived prediction score associated with same-day NS1 antigen positivity among patients with clinical suspicion of dengue in Dhaka, Bangladesh. The intended role of the score is triage support and prioritization of confirmatory testing.

## Methods and Materials

### Ethics statement

The study was approved by the Uttara Adhunik Medical College Hospital review board. Informed consent was not required because anonymized retrospective data were analyzed.

### Study design and setting

This retrospective model-development study was conducted at Uttara Adhunik Medical College Hospital, Dhaka, Bangladesh. Data were collected from patients presenting with clinical suspicion of dengue between 19 August and 12 October 2023. Patients with clinical suspicion of dengue, such as fever >37.5°C for >3 days, underwent same-day CBC and NS1 antigen testing. Same-day NS1 antigen positivity was the primary binary outcome.

### Eligibility criteria and data preparation

The initial dataset contained 743 encounters. Patients were included if they had clinical suspicion of dengue and underwent both CBC and NS1 antigen testing on the same day. To avoid correlated repeat measurements, only the earliest eligible record per patient was retained. Mean platelet volume and platelet distribution width were excluded because 115 records were missing these variables. Basophil percentage was excluded because it was uniformly recorded as 0.0 and therefore provided no discriminatory information. Records with partial or missing values in the remaining analysis variables were excluded. The final analysis dataset included 596 unique patients.

### Statistical analysis and model development

All statistical analyses were performed in R (v4.4.1) [14] using the tidyverse [15], tableone [16], glmnet [17], rms [18], and pROC [19] packages. This study was reported in accordance with relevant TRIPOD guidance for prediction model development and internal validation [20]. Descriptive characteristics were summarized overall and by NS1 status. The primary outcome was same-day NS1 antigen positivity, coded as 1 for positive and 0 for negative. Candidate predictors were derived from routinely available demographic and CBC parameters. Platelet count was rescaled per 10 x 10^9/L (Platelets10 = platelets/10). The neutrophil-to-lymphocyte ratio (NLR) was calculated as neutrophil count divided by lymphocyte count. Because of right skewness, NLR and ESR were transformed as logNLR = log(NLR) and logESR = log1p(ESR). A complete-case analysis was used for model development, and the final score was evaluated in patients with complete data for the retained predictors.

A multivariable logistic-regression framework was used. To reduce overfitting and select predictors among correlated hematological variables, we used LASSO penalized logistic regression [21], implemented with glmnet [17,22]. The candidate predictor set included age, sex, Platelets10, white blood cell count, monocyte count, hematocrit, red cell distribution width, logNLR, and logESR. The penalty parameter lambda was selected using 10-fold cross-validation. To prioritize parsimony and clinical usability, we applied the 1-standard-error rule (lambda.1se) and retained predictors with non-zero coefficients. The selected predictors were then refit in an unpenalized logistic regression model using rms::lrm [18].

### Internal validation and model performance

Discrimination was quantified using the area under the receiver operating characteristic curve (AUC). Internal validation was conducted using optimism-corrected bootstrapping with 400 resamples [23]. Calibration was assessed using a bootstrap calibration curve generated with rms::calibrate. Overall model performance and optimism-corrected indices were estimated with rms::validate, and optimism-corrected AUC was derived from Somers Dxy as AUC = (Dxy + 1)/2.

To assess incremental value beyond platelet count alone, we compared three prespecified logistic regression models using the same complete-case analysis dataset: a platelet-only model, a platelet-plus-WBC model, and the final three-variable CBC model. For each model, we estimated AUC with 95% CI, Brier score, calibration intercept, and calibration slope. 25 at the Youden-derived cutoff of 11.825 was summarized with exact binomial 95% confidence intervals, and a rounded cutoff of 12 was evaluated as a practical sensitivity analysis.

### Clinical score derivation

To improve usability, coefficients from the final model were converted into a scaled CBC-derived linear score using a standard risk-score approach [24]. Coefficients from retained predictors were scaled by the smallest absolute non-zero coefficient, excluding the intercept, and rounded to the nearest integer. Because continuous CBC values are used, the total score can take fractional values even when the weights are integers. The optimal operating point for binary classification was selected using Youden index [25] diagnostic performance at this threshold was summarized using sensitivity, specificity, positive predictive value (PPV), negative predictive value (NPV), and accuracy. The Youden-derived cutoff was selected in the apparent development dataset. Classification metrics at this cutoff should therefore be interpreted as apparent threshold performance. Risk tiers were defined using a symmetric margin of +/-10 score units around the Youden-derived cutoff.

## Results

### Study population and data flow

Of 743 initial encounters, 147 records were excluded during deduplication and quality control for missing or non-informative data. The final study population comprised 596 unique patients; 306 (51.3%) were NS1 positive and 290 (48.7%) were NS1 negative. The study population selection process is summarized in Figure 1.

**Figure 1.**
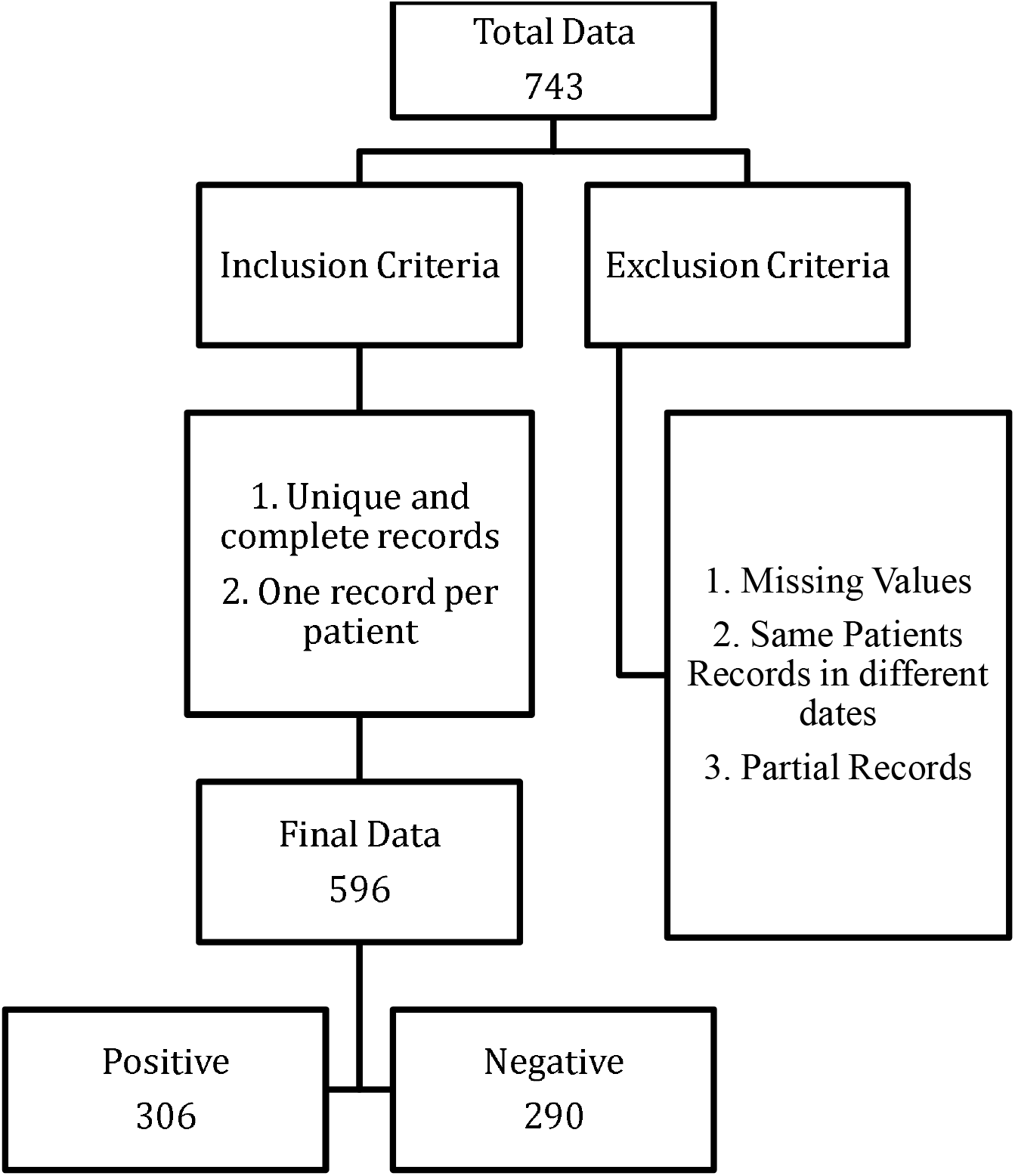
Overview of data selection criteria and study population flow.

### Baseline characteristics

The mean age was slightly higher among NS1-positive patients than NS1-negative patients, but the difference was not statistically significant (32.75 vs 30.08 years; p=0.078). Male sex was more common in the NS1-positive group (57.2% vs 45.2%; p=0.004). NS1-positive patients had lower mean WBC count, platelet count, NLR, ESR, and monocyte count than NS1-negative patients. Hematocrit did not differ significantly between groups (Table 1).

**Table 1.** Baseline characteristics of suspected dengue patients by NS1 status.

| **Variable** | **NS1 negative (n=290)** | **NS1 positive (n=306)** | **p-value** |
|---|---|---|---|
| Age, years, mean (SD) | 30.08 (19.79) | 32.75 (17.06) | 0.078 |
| Male sex, n (%) | 131 (45.2) | 175 (57.2) | 0.004 |
| WBC (10^9/L), mean (SD) | 9.29 (3.97) | 6.41 (3.68) | <0.001 |
| Platelets (10^9/L), mean (SD) | 288.16 (111.43) | 103.13 (77.36) | <0.001 |
| ESR (mm/1st hr), mean (SD) | 25.90 (23.45) | 17.60 (16.67) | <0.001 |
| NLR, mean (SD) | 2.75 (2.77) | 1.82 (2.01) | <0.001 |
| Monocytes (10^9/L), mean (SD) | 0.42 (0.21) | 0.29 (0.17) | <0.001 |

### Model development and final predictors

LASSO selection using the 1-standard-error rule retained three predictors: Platelets10, WBC count, and monocyte count. In the unpenalized refit model, all three retained predictors showed inverse associations with NS1 positivity. Platelets10 and WBC had more precise effect estimates, whereas monocyte count had a larger coefficient but a wider confidence interval (Table 2).

**Table 2.** Final multivariable logistic regression model.

| **Variable** | **Beta coefficient** | **Odds ratio** | **95% confidence interval** |
|---|---|---|---|
| Intercept | 5.376 | 216.254 | 88.477-528.567 |
| Platelets10 | -0.228 | 0.796 | 0.767-0.826 |
| White blood cells (10^9/L) | -0.101 | 0.904 | 0.823-0.993 |
| Monocytes (10^9/L) | -1.534 | 0.216 | 0.032-1.473 |

The final logistic model was:
logit(p) = 5.376453 − 0.2278 × Platelets10 − 0.10084 × WBC − 1.53414 × Monocyte
where p is the predicted probability of NS1 antigen positivity, Platelets10 = platelet count/10, WBC is white blood cell count, and Monocyte is monocyte count.
The predicted probability can be calculated as:
p = 1 / [1 + exp(−LP)]

### Model performance and internal validation

The final model showed strong discrimination. The apparent AUC was 0.9436, and bootstrap internal validation with 400 resamples yielded an optimism-corrected AUC of 0.9429. The bootstrap calibration plot showed good overall agreement between predicted and observed NS1 positivity, with minor miscalibration in the middle range of predicted probabilities. The mean absolute error between predicted and observed probabilities was 0.057. The ROC curve and calibration plot are shown in Figure 2.

**Figure 2.**
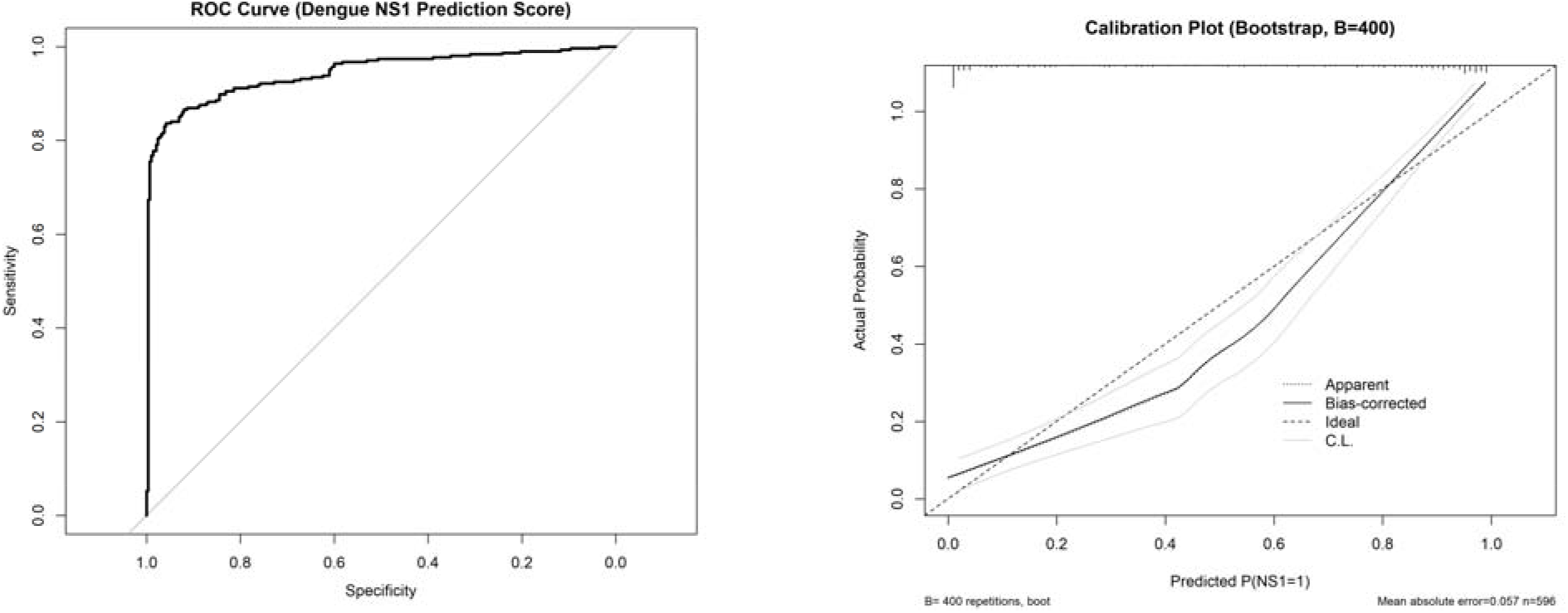
ROC curve and bootstrap calibration plot for the final CBC-derived score. (A) Receiver operating characteristic curve. (B) Bootstrap calibration plot based on 400 resamples. The final score is intended to estimate NS1 antigen positivity in this dataset, not to replace confirmatory testing.

To assess whether the final three-variable score added value beyond platelet count alone, we compared the final model with two simpler benchmark models using the same analysis dataset (n=596; 306 NS1 positive and 290 NS1 negative). The platelet-only model had an AUC of 0.931 (95% CI, 0.911-0.952) and a Brier score of 0.102. Adding WBC improved discrimination and reduced prediction error (AUC, 0.943; 95% CI, 0.924-0.962; Brier score, 0.093). The final CBC score model had an AUC of 0.944 (95% CI, 0.925-0.963) and a Brier score of 0.092, representing a modest improvement over platelet count alone (Delta AUC, 0.012; DeLong p=0.00364) (Table 3).

**Table 3.** Benchmark model comparison using the same complete-case dataset.

| **Model** | **N** | **NS1+** | **NS1-** | **AUC** | **95% CI** | **Brier** | **Delta AUC** | **DeLong p** | **Calibration intercept** | **Calibration slope** |
|---|---|---|---|---|---|---|---|---|---|---|
| Platelet-only model | 596 | 306 | 290 | 0.931 | 0.911-0.952 | 0.102 |  |  | 0.000 | 1.000 |
| Platelet + WBC model | 596 | 306 | 290 | 0.943 | 0.924-0.962 | 0.093 | 0.011 | 0.00395 | 0.000 | 1.000 |
| Final CBC score model | 596 | 306 | 290 | 0.944 | 0.925-0.963 | 0.092 | 0.012 | 0.00364 | 0.000 | 1.000 |

At the Youden-derived cutoff of 11.825, the final integer score had an AUC of 0.944 (95% CI, 0.925-0.963), sensitivity of 0.833 (95% CI, 0.787-0.873), specificity of 0.966 (95% CI, 0.938-0.983), PPV of 0.962 (95% CI, 0.932-0.982), NPV of 0.846 (95% CI, 0.802-0.883), and accuracy of 0.898 (95% CI, 0.870-0.921) (Table 5).

The rounded cutoff of 12 produced similar performance to the Youden-derived cutoff of 11.825, with sensitivity 0.824, specificity 0.966, PPV 0.962, NPV 0.838, and accuracy 0.893 (Table 6).

Risk-tier analysis showed marked separation of observed NS1 positivity across score categories. In the high-risk tier (score >21.825), 181 of 182 patients (99.5%) were NS1 positive. The intermediate-risk tier included 145 patients with an observed positivity rate of 66.2%, while the low-risk tier included 269 patients with a positivity rate of 10.8%. Because the cutoff and tier boundaries were derived in the same dataset, these threshold-based estimates may be optimistic and require confirmation in independent data (Table 7).

### CBC-derived score and diagnostic performance

The CBC score used integer weights derived from the final model coefficients: intercept +53 points, Platelets10 -2 points, WBC -1 point, and monocyte count -15 points (Table 4). The score was calculated as follows:

**Score = 53 -2 x Platelets10 - 1 x WBC - 15 x Monocyte**

**Table 4.** CBC-derived prediction score: scaled weights for retained predictors.

| **Variable** | **Beta** | **Points** |
|---|---|---|
| (Intercept) | 5.376453 | 53 |
| Platelets10 | -0.227798 | -2 |
| WhiteBloodCells(109L) | -0.100843 | -1 |
| Monocyte(109L) | -1.534143 | -15 |

**Table 5.** Diagnostic performance of the final integer score at the Youden-derived cutoff.

| **Metric** | **Estimate** | **95% CI** | **CI method** |
|---|---|---|---|
| AUC | 0.944 | 0.925-0.963 | DeLong CI for AUC |
| Sensitivity | 0.833 | 0.787-0.873 | Exact binomial CI |
| Specificity | 0.966 | 0.938-0.983 | Exact binomial CI |
| PPV | 0.962 | 0.932-0.982 | Exact binomial CI |
| NPV | 0.846 | 0.802-0.883 | Exact binomial CI |
| Accuracy | 0.898 | 0.870-0.921 | Exact binomial CI |

**Table 6.**
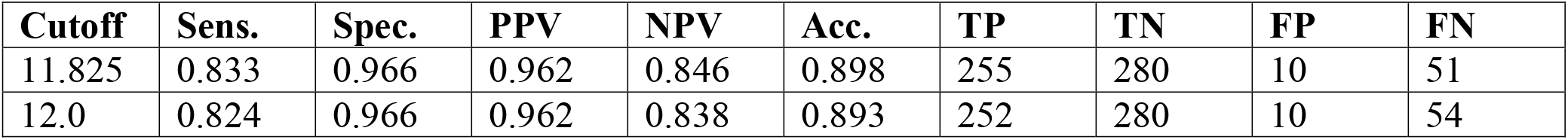
Practical cutoff sensitivity analysis.

**Table 7.** Observed NS1 positivity by risk tier.

| **Risk tier** | **High** | **Intermediate** | **Low** |
|---|---|---|---|
| Number | 182 | 145 | 269 |
| NS1 positive | 181 | 96 | 29 |
| NS1 negative | 1 | 49 | 240 |
| Observed positivity rate | 0.995 | 0.662 | 0.108 |

**Table 8.** Subgroup discrimination performance by age category.

| **Age group** | **Adult** | **Pediatric** |
|---|---|---|
| N | 467 | 129 |
| NS1 positive | 252 | 54 |
| AUC | 0.945 | 0.939 |

## Discussion

In this retrospective single-center study of suspected dengue patients in Dhaka, we developed and internally validated a parsimonious CBC-derived score associated with same-day NS1 antigen positivity. The final model used only three routinely available parameters: Platelets10, WBC count, and monocyte count. The model demonstrated strong internal discrimination, with an optimism-corrected AUC of 0.9429, and performance was similar in adult and pediatric subgroups. These findings suggest that routine CBC patterns contain substantial information about the likelihood of NS1 antigen positivity during the study period.

The selected predictors are consistent with existing evidence. Leukopenia and thrombocytopenia have been reported as useful early markers of dengue infection during outbreak settings [11]. The Early Dengue Classifier developed by Tuan and colleagues incorporated WBC and platelet count among its predictors [12]. Poottasane and colleagues also reported a CBC-based NS1-positivity score that included WBC and platelet parameters, although their score used monocyte distribution width rather than monocyte count [13]. More complex decision-support approaches have also been evaluated; for example, Sa-ngamuang and colleagues compared clinician diagnosis with Bayesian network models using dengue clinical data with and without NS1 rapid test results [26]. Together, these studies support the relevance of CBC-derived signals for early dengue triage, while also emphasizing that prediction models require local and external validation before implementation.

The benchmark analysis was included because platelet count is a strong and clinically familiar marker in suspected dengue. Comparing the final model with platelet-only and platelet-plus-WBC models helps clarify the incremental value of the final CBC-derived score. The final model showed a modest AUC improvement over platelet count alone and a slightly lower Brier score, but these findings should be interpreted as internal evidence only. The score should be used, if at all, as a triage adjunct for NS1 antigen positivity rather than a definitive CBC-derived NS1-positivity triage score. The rounded cutoff analysis supports practical usability because a cutoff of 12 performed similarly to 11.825, but both thresholds require confirmation in independent datasets before routine clinical use.

The direction of association in the final model is clinically plausible. Thrombocytopenia and leukopenia are common hematologic features in dengue and are frequently used as supportive evidence during clinical assessment [1,8,27]. Monocyte involvement is also biologically plausible because experimental and clinical studies suggest that monocytes can be susceptible to dengue virus infection and may contribute to viral replication and immune response dynamics [28,29]. However, this study was designed for prediction rather than causal inference. The monocyte coefficient had a wide confidence interval; therefore, the monocyte component should be interpreted cautiously and examined in external validation.

The practical advantage of the score is that it relies only on CBC parameters that are inexpensive and widely available. In high-volume settings where NS1 testing may be delayed or capacity-limited, the score may help prioritize confirmatory testing or closer follow-up among patients with clinical suspicion of dengue. Patients in the high-risk tier had a high observed probability of NS1 positivity in this dataset and could be prioritized for confirmatory testing and follow-up if clinically appropriate. Intermediate-risk results should be treated as an uncertainty zone. Low-risk results should not be used to rule out dengue when clinical suspicion remains high, because NS1 test performance and CBC patterns vary with illness timing and other factors [9,10,30-32].

### Limitations and future directions

Several limitations should be acknowledged. First, the study used retrospective data from a single center, which may limit transportability to other hospitals, regions, seasons, and dengue prevalence settings. Second, the outcome was same-day NS1 antigen positivity rather than dengue infection confirmed by a broader reference standard such as RT-PCR, paired serology, or expert adjudication. Third, all predictors were CBC-derived, and no illness-day, symptom, serotype, secondary-infection, severity, or comorbidity variables were available. Fourth, although bootstrap internal validation was performed, the Youden cutoff and risk-tier boundaries were derived in the same dataset; threshold-based sensitivity, specificity, PPV, NPV, accuracy, and tier positivity rates may therefore be optimistic. Finally, the final model showed only modest incremental improvement over platelet count alone, reinforcing that the score should be viewed as a formalized CBC-only triage adjunct rather than a replacement for confirmatory testing. Future work should externally validate the score in independent Bangladeshi cohorts and assess whether it improves triage workflow in prospective use.

### Conclusion

A simple CBC-derived score using platelet count, WBC count, and monocyte count was strongly associated with same-day dengue NS1 antigen positivity in this retrospective cohort of suspected dengue patients in Dhaka. Benchmark analysis showed that the final model improved modestly over platelet count alone. The score may support CBC-only triage and prioritization of confirmatory testing during high-volume periods, but it does not diagnose dengue and should not replace NS1, RT-PCR, serology, or clinical judgment. External validation is required before routine clinical use.

## Data Availability

All data produced in the present study are available upon reasonable request to the corresponding author.

## Authors’ Contributions

Rasel Khan Nishan: Conceptualization, methodology, formal analysis, visualization, and writing – original draft; Md. Shaha Alam: Investigation, resources, data curation, and writing – review and editing; Abul Kalam Azad: Investigation, data curation, and writing – review and editing; Md. Murshed Hasan Sarkar: Methodology, validation, and writing – review and editing; Tanjum Ahmed Nodee: Data curation, validation, and writing – review and editing; Md. Zakaria Mia: Investigation, data curation, and writing – review and editing; Md. Ibrahim Miah: Methodology, validation, and writing – review and editing; Md. Shariful Islam: Conceptualization, supervision, project administration, and writing – review and editing. All authors reviewed and approved the final version of the manuscript and agree to be accountable for their respective contributions to the work.

